# Not So Fast, I’m Serofast: Using Innovative Education Techniques to Drive Management of People with Syphilis

**DOI:** 10.1101/2025.07.24.25330660

**Authors:** Dennis Kan, Molly Stenzel, Julie Thompson, Jennie De Gagne, Kara McGee

## Abstract

**Background:** An increase in syphilis cases in the United States and the global shortage of Benzathine Penicillin G (BPG) calls for evidence-based optimization. Confusing serologies and incomplete sexual history exacerbate the overprescription of BPG, especially for patients with “serofast” serologies. Seronegative status develops through natural decline and not through additional therapy.

**Method:** A longitudinal pre-post design was implemented within a health maintenance organization serving Oregon and Southwest Washington. Interventions included microlearning education, allowing quick access to clinical recommendations and actionable patient education empowers patients.

**Results:** BPG was administered 187 times (69 pre-intervention, 118 post-intervention), and inappropriate administration showed a 7.9% relative decline (10.1% to 9.3%). There was no difference in outcomes between the newly infected and reinfected patients. Patient education utilization increased from 4.3% to 6.8%, representing a 58.1% relative improvement. Clinician microlearning utilization decreased by 25.3% during the post-intervention period.

**Conclusion:** This project demonstrated modest improvements in the administration of appropriate BPG and utilization of patient education, although the results did not reach statistical significance. Future interventions should focus on automating patient education and enhancing the visibility of clinical resources. This project highlighted the importance of obtaining patients’ treatment histories and recent exposure risks. The increasing prevalence of syphilis among women who have sex with men (WSM) represents an epidemiological shift with implications for maternal-child health. The growing prevalence of syphilis among non-traditional risk groups emphasizes the importance of comprehensive sexual health screening approaches to mitigate adverse outcomes.

**What is already known on this topic:** Syphilis is a sexually transmitted infection (STI) with public health urgency and a global impact due to shortage and emergence. There are studies on the effects of syphilis and public health, but they do not address how to combat the syphilis epidemic. A common practice is that many medical providers are not familiar with how to interpret syphilis results, which can lead to overtreatment of patients, especially those who are serofast. There are also limited studies that look at the feasibility and utilization of using education to lower syphilis rates.

**What this study adds:** From this study, we found that using innovative educational methods, microlearning, and patient-centered patient education can modestly reduce the inappropriate treatment of syphilis. Syphilis treatment is complex, and knowledge gaps that exist include too frequent testing and not obtaining an adequate sexual health history. This study also confirms that there is an importance of screening other populations and not just men who have sex with men (MSM).

**How might this study affect research, practice, or policy?:** This study shows the urgency of the treatment shortage and the rising rate of syphilis, calling for an innovative approach. Institutions need to look at other ways to address the syphilis epidemic, such as utilizing practical tools from this study to decrease the inappropriate use of syphilis treatment. These interventions are helpful for medical providers and provide patients with a means to advocate for their health. Refocusing on the clinical need of appropriate history taking in conjunction with serologies to properly treat people with syphilis. Other innovation measures, such as microlearning videos and artificial intelligence, can be tools to aid in appropriate syphilis treatment.

## Introduction

Syphilis is a sexually transmitted infection (STI) that has increased by 80% in the United States over the past 5 years. Alongside this increase, a global shortage of raw materials has led to a scarcity of Benzathine Penicillin G (BPG) injections used to treat syphilis, resulting in adverse outcomes, including increasing cases of congenital syphilis.

Men who have sex with men (MSM), people with human immunodeficiency virus (PWH), and women are disproportionately affected by syphilis. MSM accounts for 45% of all men with primary and secondary (P&S) syphilis infections, and 36.4% of the MSM with syphilis are PLW. The United States has seen a 19.2 % increase in primary and secondary (P&S) syphilis infections in women from 2021 to 2022 in 36 states and the District of Columbia [5].

Understanding syphilis titers coupled with sexual health history is imperative for appropriate treatment. Serological non-responders, or "serofast," refer to persistent low non-treponemal titers after a 4-fold decrease and resolution of symptoms [6, 7]. Overtreatment of patients with serofast syphilis titers with BPG has no additional benefits, as seronegative titers occur from natural decline [8, 9].

Patient education often omits essential information regarding syphilis, such as side effects of BPG treatment. In addition, incomplete sexual history leads to overprescription of BPG, particularly in patients with serofast serologies [9]. The current BPG shortage is an emerging global issue exacerbated by the overtreatment of patients with syphilis serofast serologies.

This project seeks to educate clinicians and patients on syphilis serologies and indications for BPG. Microlearning has emerged as a practical approach to delivering “bite-sized” information in clinical settings, thereby boosting confidence and facilitating efficient knowledge acquisition among clinical learners. Patient education is equally important with intrinsic values in advocating for their health, and delivering actionable information helps empower them to make informed decisions [12]. One practical approach is to include educational material in the after visit summary (AVS), a vital information source for patients after a BPG injection [13]. Together, innovative strategies such as microlearning and patient-facing education can empower clinicians to be effective stewards of BPG, enabling patients to advocate for their care, ultimately improving outcomes and reducing inappropriate treatment.

This project aimed to reduce unindicated BPG administration by 20% each month within 3 months, while maintaining or improving appropriate BPG administration in people diagnosed with syphilis during the post-intervention period. In addition, increase attaching patient education to an after-visit summary (AVS) and utilization of the clinician’s syphilis practice resource by 20% each month post-intervention. This project focused on using BPG for syphilis and not as an alternative agent due to patient allergies or the treatment of neurosyphilis, ocular syphilis, or otosyphilis, as the recommended treatment omits BPG.

## Materials and Methods

A longitudinal pre-post design was employed from October 2024 to December 2024 (pre-intervention) and from January 2025 to March 2025 (post-intervention) within a health maintenance organization (HMO). This HMO serves the State of Oregon and Southwest Washington, comprising 31 ambulatory care facilities and two tertiary care hospitals (Northwest Permanente, 2024). The implementation focused on ambulatory settings, including primary care, Urgent Care (UC), and Nurse Treatment Rooms (NTR). NTR, clinics run by registered nurses (RNs), provide patients with antibiotic injections. Although these interventions are primarily ambulatory, they are also available in the Emergency Department (ED).

### Patient and Public Involvement

Interviews were conducted with key stakeholders to help inform the development of modifications to existing patient education and clinician practice guidelines. These three key stakeholders, which included a Nurse Practitioner (NP), an RN, and a patient who received syphilis treatment, were selected and interviewed based on their interaction with the site where the intervention took place. The interview gathered information from stakeholders, including barriers to current education and workflow processes, among other topics. For example, information was added to the patient education to help forecast receiving a call from public health for contact tracing. The three interviews with key stakeholders are available in Appendix #1. Results and findings from this project were later made available to clinic staff and other stakeholders within the organization, including the NP and RN.

### Interventions

Two interventions used in this project: actionable education to facilitate patients’ health advocacy, and clinician microlearning to empower clinicians to feel confident in managing syphilis. In addition to interviewing end users, a literature search was performed to identify innovative educational strategies for both clinicians and patients (Appendix #2).

#### Clinician Practice Resource Microlearning

Clinicians utilize the organization’s clinical library as a practice resource to help guide patient management. The syphilis screening guidance was updated to include a concise summary of the guidelines, succinct algorithms for syphilis screening, an infographic to aid in the interpretation of Rapid Plasma Reagin (RPR) results, and contact information for local public health (Appendix #3). These resources were designed for quick reference by prescribing clinicians on busy clinic days.

#### Actionable Patient Education

The patient education components were updated to include topics such as the Jarisch-Herxheimer (J-H) reaction, ways to treat possible side effects, what "serofast" meant, actions to take if they became re-exposed to syphilis, and the importance of letting their provider know about their history of syphilis (Appendix #4). The patients were monitored for 30 min after BPG was administered in NTR. During the monitoring period, RNs reviewed the "talking points" with the patient and bullet-pointed items within the new patient education. Patient education is also attached to the AVS.

### Data Collection

Data were collected using the organization’s electronic health record (EHR) reporting functionality. BPG administration and reactive RPR reports were collected monthly during the pre- and post-intervention period.

Patient data collected included sex identity, sexual orientation (when available), HIV status, syphilis infection and treatment history, patient risk factors, and whether syphilis education was attached to the AVS. The use of BPG for indications other than syphilis treatment and patients allergic to penicillin was excluded. The criteria for the appropriateness of BPG treatment were based on syphilis staging (Appendix #5) and the World Health Organization (WHO) syphilis treatment guidelines [14]. The clinical library administrator provided raw counts of the number of times the syphilis screening resource page was accessed, which were collected monthly during the pre- and post-intervention periods.

### Statistical Analysis

Fisher’s exact statistical method was used to compare the appropriate administration of BPG among people who received it, specifically examining the percentage of unindicated administration before and after the intervention. Descriptive statistics were used to analyze the total number of patients with reactive RPR who met the BPG treatment criteria. Fisher’s exact test was used to compare the pre- and post-intervention percentages of syphilis education attached to a patient’s AVS. Finally, a run chart displaying the times the clinician’s syphilis resource was accessed during the pre- and post-intervention periods. IBM SPSS version 29 was used for the statistical analysis, with the alpha set to 0.05

## Results

### Patient Demographics When Receiving BPG

BPG was administered 187 times (69 pre-intervention and 118 post-intervention) to 112 unique patients (48 patients pre-intervention and 64 patients post-intervention). Several demographic variables of the patients were compared when they received BPG. The distribution of gender identity did not differ significantly between the groups (χ² [4, N = 187] = 2.53, p = .638). The most common gender identity was male, comprising 66.7% in the pre-intervention group and 62.7% in the post-intervention group. HIV status was also similar across groups (χ² [1, N = 187] = 0.63, p = .429), with the majority of both groups testing negative (84.1% vs. 88.1%). In contrast, a significant difference was observed in sexual activity categories (χ² [7, N = 187] = 14.92, p = .037). The most common sexual activity reported was WSM (women who had sex with men), with 29.0% in the pre-intervention group and 33.1% in the post-intervention group. However, there were more patients in post-intervention whose sexual orientation was not documented, compared to the pre-intervention (10.1% vs. 27.1%). Lastly, syphilis staging significantly differed between the groups (χ² [8, N = 187] = 20, p = .012). The most frequently reported category in the post-intervention group was Late Latent syphilis (62.7%) compared to the pre-intervention group (44.9%). Table 1 shows their demographic characteristics.

**Table 1:**
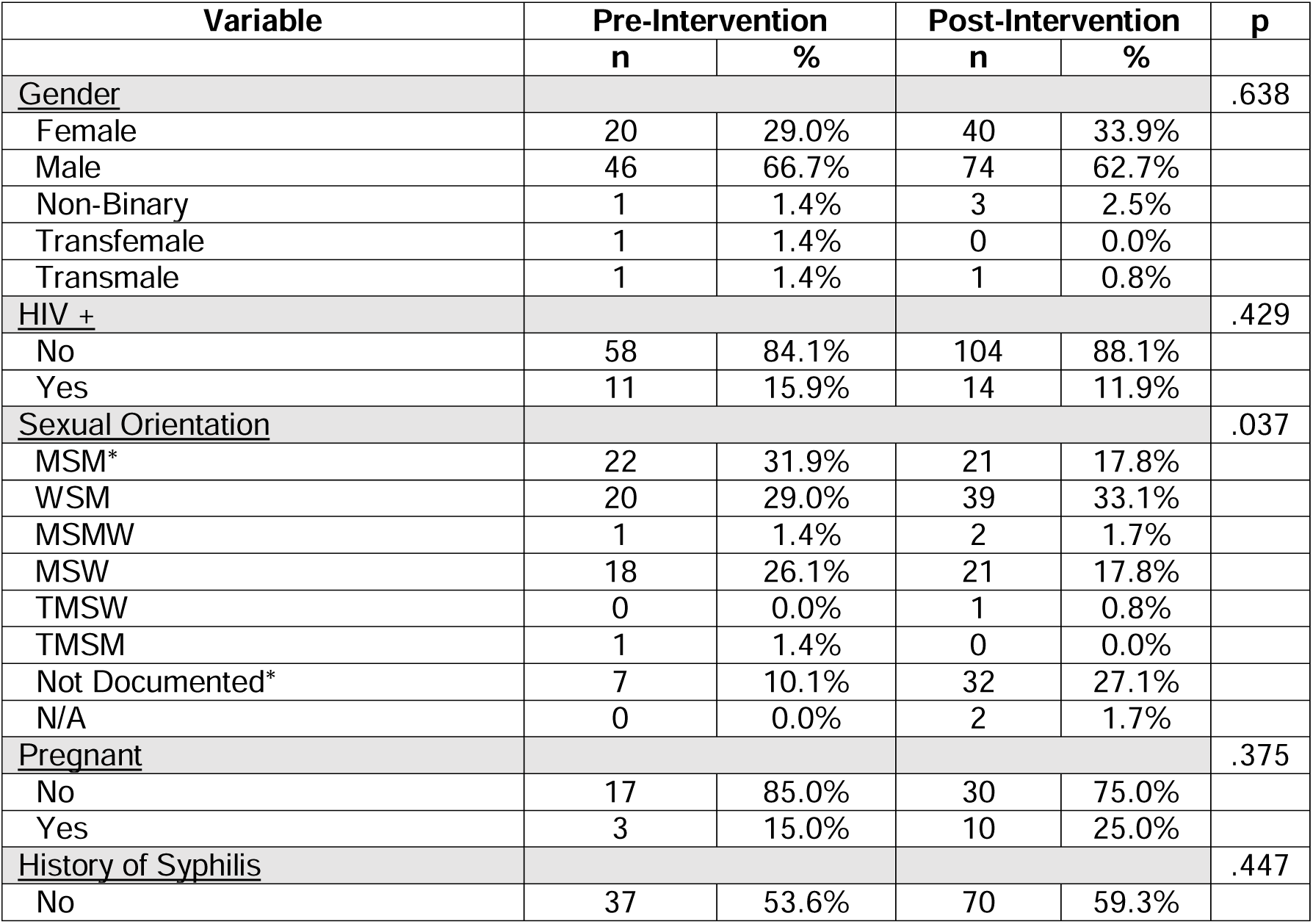
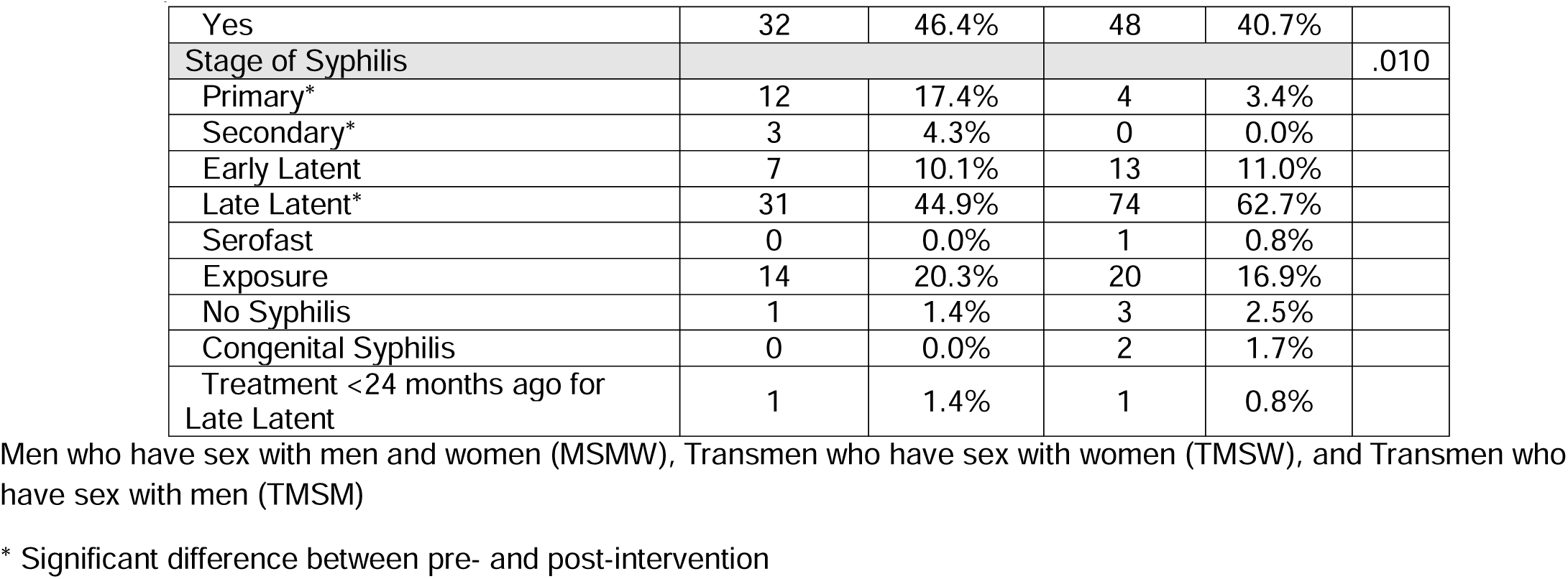
Patient Demographics Per Occurrence of BPG Injection:

| Variable | Pre-Intervention |  | Post-Intervention |  | p |
| --- | --- | --- | --- | --- | --- |
|  | n | % | n | % |  |
| **Gender** |  |  |  |  | .638 |
| Female | 20 | 29.0% | 40 | 33.9% |  |
| Male | 46 | 66.7% | 74 | 62.7% |  |
| Non-Binary | 1 | 1.4% | 3 | 2.5% |  |
| Transfemale | 1 | 1.4% | 0 | 0.0% |  |
| Transmale | 1 | 1.4% | 1 | 0.8% |  |
| **HIV +** |  |  |  |  | .429 |
| No | 58 | 84.1% | 104 | 88.1% |  |
| Yes | 11 | 15.9% | 14 | 11.9% |  |
| **Sexual Orientation** |  |  |  |  | .037 |
| MSM* | 22 | 31.9% | 21 | 17.8% |  |
| WSM | 20 | 29.0% | 39 | 33.1% |  |
| MSMW | 1 | 1.4% | 2 | 1.7% |  |
| MSW | 18 | 26.1% | 21 | 17.8% |  |
| TMSW | 0 | 0.0% | 1 | 0.8% |  |
| TMSM | 1 | 1.4% | 0 | 0.0% |  |
| Not Documented* | 7 | 10.1% | 32 | 27.1% |  |
| N/A | 0 | 0.0% | 2 | 1.7% |  |
| **Pregnant** |  |  |  |  | .375 |
| No | 17 | 85.0% | 30 | 75.0% |  |
| Yes | 3 | 15.0% | 10 | 25.0% |  |
| **History of Syphilis** |  |  |  |  | .447 |
| No | 37 | 53.6% | 70 | 59.3% |  |
| Yes | 32 | 46.4% | 48 | 40.7% |  |
| Stage of Syphilis |  |  |  |  | .010 |
| Primary* | 12 | 17.4% | 4 | 3.4% |  |
| Secondary* | 3 | 4.3% | 0 | 0.0% |  |
| Early Latent | 7 | 10.1% | 13 | 11.0% |  |
| Late Latent* | 31 | 44.9% | 74 | 62.7% |  |
| Serofast | 0 | 0.0% | 1 | 0.8% |  |
| Exposure | 14 | 20.3% | 20 | 16.9% |  |
| No Syphilis | 1 | 1.4% | 3 | 2.5% |  |
| Congenital Syphilis | 0 | 0.0% | 2 | 1.7% |  |
| Treatment <24 months ago for Late Latent | 1 | 1.4% | 1 | 0.8% |  |

Men who have sex with men and women (MSMW), Transmen who have sex with women (TMSW), and Transmen who have sex with men (TMSM)
\* Significant difference between pre- and post-intervention

A chi-square test of independence examined the distribution of sexual activity categories between pre-implementation and post-implementation after removing “N/A” and “Not Documented” patients from the samples. The most common sexual orientations in the pre-intervention were MSM (35.5%) and WSM (32.3%). Similarly, at post-intervention, WSM was the most common (46.4%), followed by MSM (25.0%) and men who had sex with women (MSW) (25.0%). The overall distribution of sexual activity categories did not differ significantly between the pre-intervention and post-intervention groups (χ² [5, N = 146] = 5.52, p = .356).

### Patient Demographics of Reactive RPRs

A total of 404 reactive RPR test results were obtained from 297 unique patients (170 pre-intervention and 187 post-intervention). Chi-square analysis was conducted to evaluate the demographic characteristics of each RPR sample. The most common gender identity in both groups was male, with 145 reactive RPR results (75.5%) in the pre-intervention period and 170 reactive RPR results (80.2%) in the post-intervention period. Female patients comprised 34 reactive RPR results (17.7%) in pre-intervention and 37 reactive RPR results (17.5%) in post-intervention. The chi-square test revealed no statistically significant differences in gender identity distributions between the groups (χ² (4) = 4.70, p = .319). Of the 404 reactive RPR samples, 172 belonged to patients with a history of syphilis pre-intervention, and 184 belonged to patients with a history of syphilis post-intervention, χ² (1) = 0.75, p = .386. Serofast was the most common stage, based on the patient’s reactive RPR, with 76.0% in the pre-intervention group (n = 146) and 74.1% in the post-intervention group (n = 157). This was followed by late latent (8.3% vs. 9.0%), and early latent (5.2% vs. 8.0%). Other less frequent classifications were exposure (3.6% vs. 1.9%), primary syphilis (2.6% vs. 0.9%), secondary syphilis (0.5% vs. 0.5%), congenital syphilis (0.0% vs. 0.5%), and no syphilis (0.0% vs 0.5%). The distribution of syphilis stages was similar among all the patients (χ² (9) = 13.39, p = .146) (Table 2).

**Table 2:** Patient Demographics Per Reactive RPR Sample:

| Variable | Pre-Intervention |  | Post-Intervention |  | p |
| --- | --- | --- | --- | --- | --- |
|  | n | % | n | % |  |
| **Gender** |  |  |  |  | .319 |
| Female | 34 | 17.7% | 37 | 17.5% |  |
| Male | 145 | 75.5% | 170 | 80.2% |  |
| Non-Binary | 5 | 2.6% | 2 | 0.9% |  |
| Transfemale | 3 | 1.6% | 1 | 0.5% |  |
| Transmale | 5 | 2.6% | 2 | 0.9% |  |
| **History of Syphilis** |  |  |  |  | .387 |
| No | 20 | 10.4% | 28 | 13.2% |  |
| Yes | 172 | 89.6% | 184 | 86.8% |  |
| **Stage of Syphilis** |  |  |  |  | .146 |
| Primary | 5 | 2.6% | 2 | 0.9% |  |
| Secondary | 1 | 0.5% | 1 | 0.5% |  |
| Early Latent | 10 | 5.2% | 17 | 8.0% |  |
| Late Latent | 16 | 8.3% | 19 | 9.0% |  |
| Exposure | 7 | 3.6% | 4 | 1.9% |  |
| Serofast | 146 | 76.0% | 157 | 74.1% |  |
| Treatment <12 months ago for P&S or Early Latent | 4 | 2.1% | 0 | 0.0% |  |
| Treatment <24 months ago for Late Latent | 3 | 1.6% | 10 | 4.7% |  |
| Congenital Syphilis | 0 | 0.0% | 1 | 0.5% |  |
| No Syphilis | 0 | 0.0% | 1* | 0.5% |  |

\*Patient had a 1:1 RPR titer, followed by a non-reactive RPR titer the next day

### Appropriateness of BPG administration

Post-intervention, inappropriate BPG administration decreased by 9.1% between January and February and increased by 10.6% in March (Figure 1). Fisher’s exact test was used to examine the association between the intervention groups and whether BPG was ordered appropriately. The results indicated no statistically significant difference between the groups (p = .854). Most patients in the pre-intervention (89.9%, 62 of 69) and post-intervention (90.7%, 107 of 118) received or had BPG appropriately ordered. Inappropriate administration declined from 10.1% overall in pre-intervention to 9.3% overall post-intervention, a 7.9% relative decline.

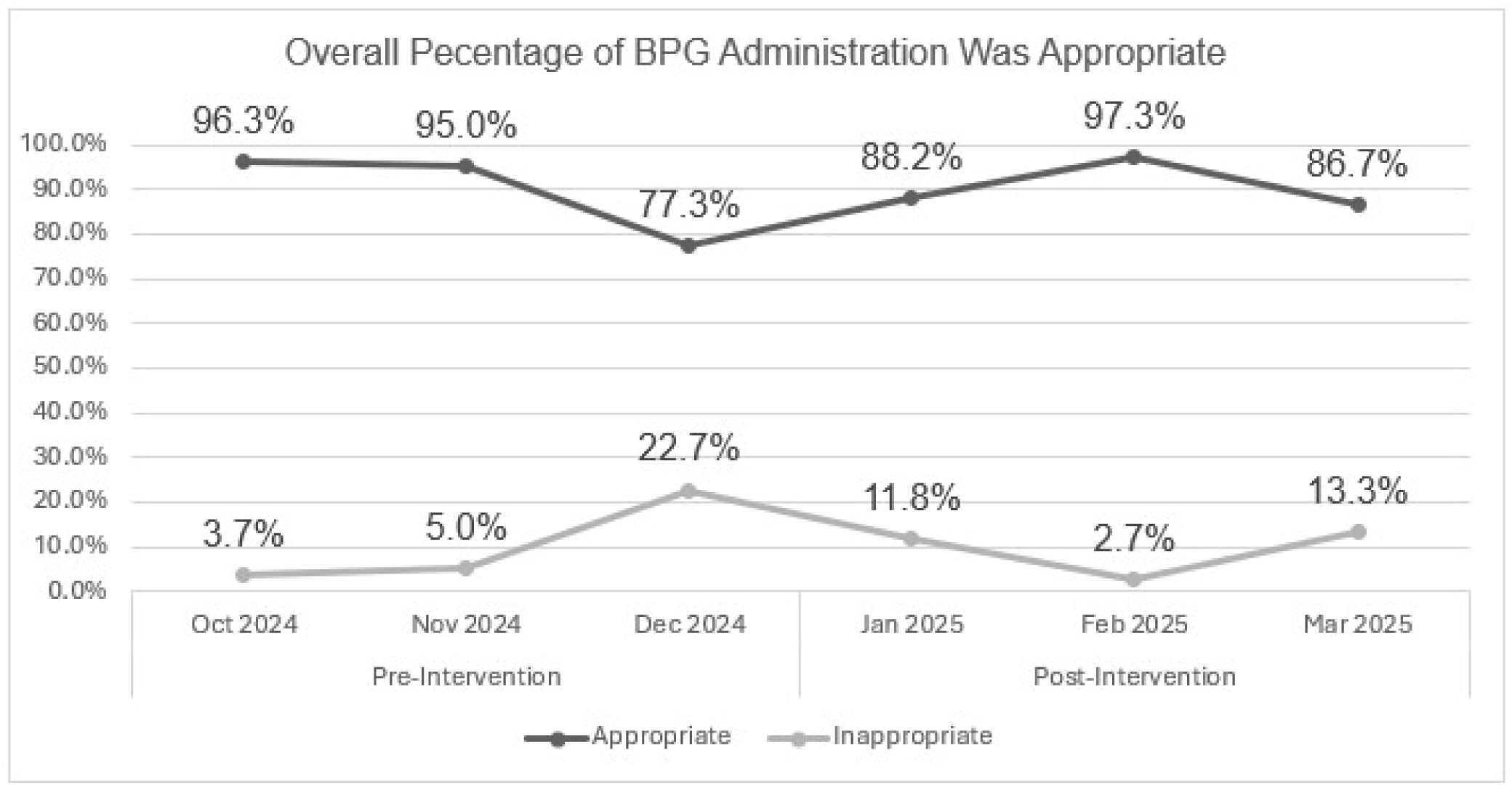

### BPG administration among people with syphilis

Fisher’s exact test was used to examine the association between the reactive RPR intervention groups and whether BPG was ordered appropriately. The results indicated no statistically significant difference between the groups (p = .999). Most patients in the pre-intervention group (94.7%, 36 of 38) and the post-intervention group (93.3%, 42 of 45) received or had BPG appropriately ordered (Figure 2). When comparing patients without a history of syphilis who had a reactive RPR, inappropriate BPG orders declined from pre-intervention (n = 1, 5%) to post-intervention (n = 1, 3.7%). For those with a history of syphilis, inappropriate BPG orders also declined from pre-intervention (n = 3, 15%) to post-intervention (n = 2, 11.1%), although both groups were not significant, p = .999.

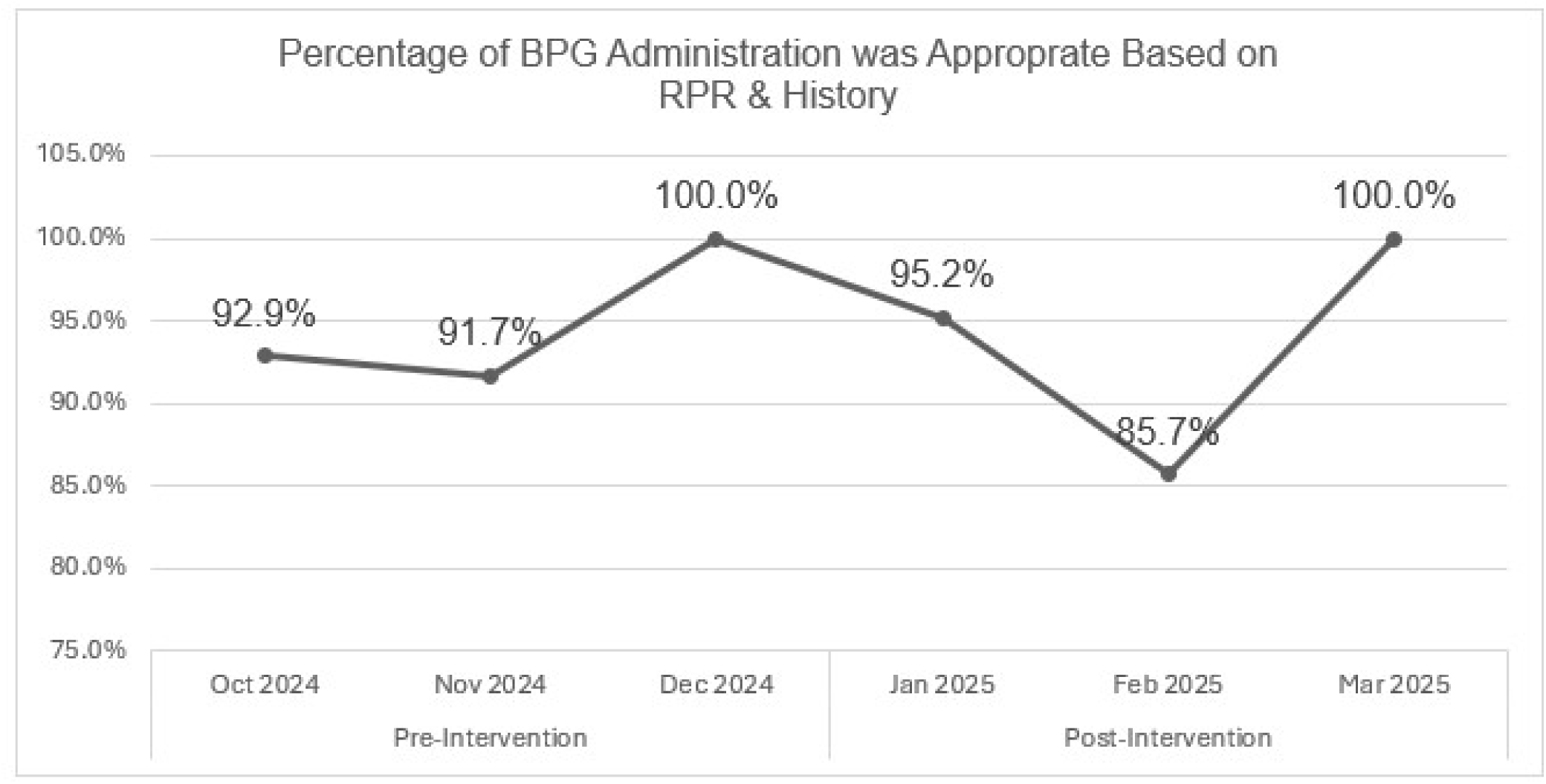

### Attachment of patient education to an AVS after a visit for a BPG injection

At least 5% of encounters with patient education attached to the AVS were recorded in all three implementation months. Fisher’s exact test was used to assess the relationship between the intervention group and whether patient education was associated with AVS. A total of 187 BPG administration events were included, comprising 69 events in the pre-intervention period and 118 in the post-intervention period. Although attachment rates increased from pre-intervention (4.3%, 3 of 69) to post-intervention (6.8%, 8 of 118), demonstrating a relative improvement of 58.1%, it was not statistically significant (p = .749) (Figure 3).

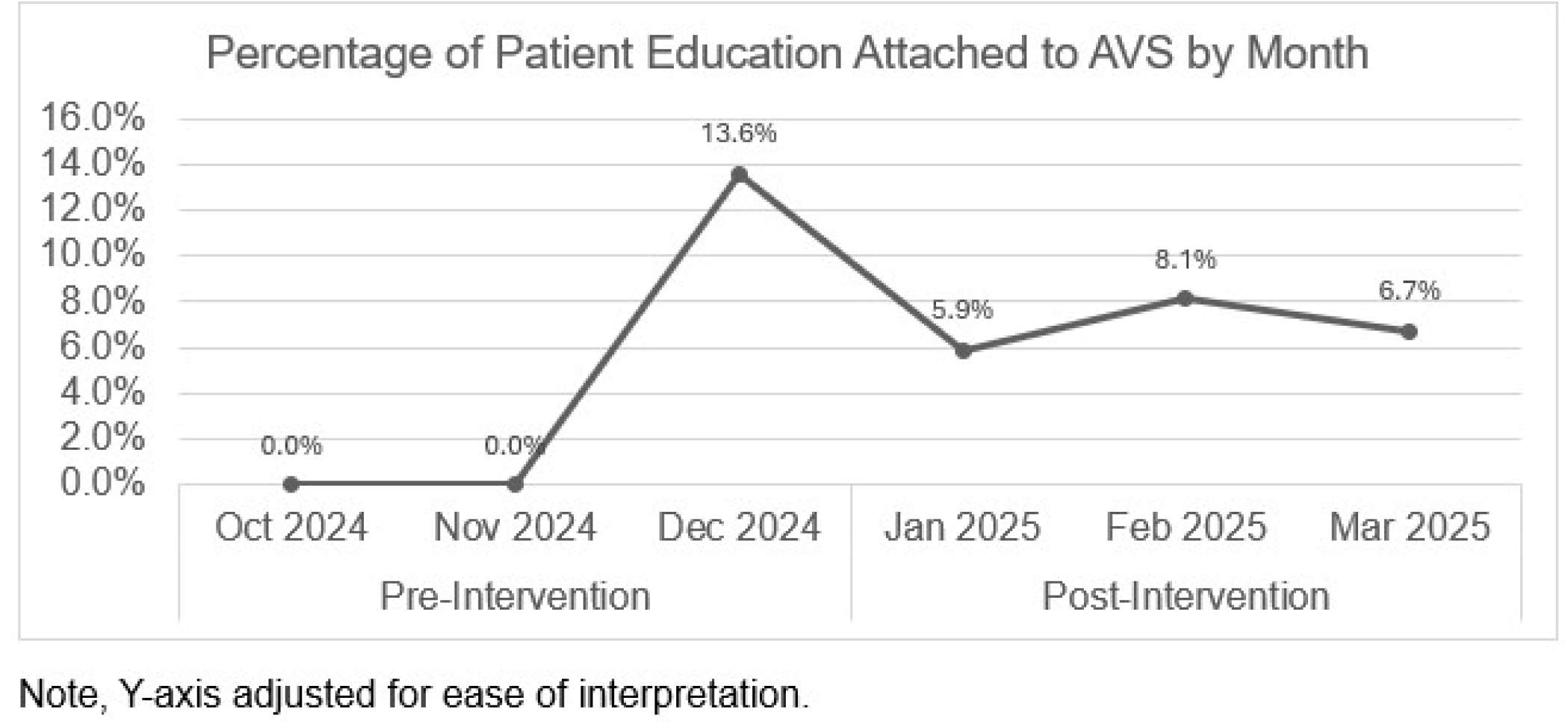

### Utilization of syphilis practice resource microlearning within the organization’s clinical library

The syphilis clinician practice resource microlearning was viewed 83 times in the pre-intervention period and 62 times in the post-intervention period, representing a 25.3% decrease.

## Discussion

This study demonstrated a relative decrease in the inappropriate administration of BPG, maintenance of appropriate BPG administration for people with newly acquired syphilis, and increased use of patient education materials. The project did not increase the utilization of clinician learning resources for syphilis diagnosis.

Improvements in the clinical management of syphilis have been noted, particularly in the reduction of inappropriate BPG administration. No significant difference in BPG administration was observed among patients with reactive RPR, indicating that educational interventions did not result in additional adverse effects. These findings align with the WHO guidelines, which emphasize the importance of correct BPG use as first-line treatment for syphilis [14].

Notably, there was an increase in the number of patients with WSM who received BPG in the post-intervention group. This increase coincided with an increase in pregnant individuals in the post-intervention group (n = 3, 15.0% vs. n = 10, 25.6%). The Oregon Health Authority (OHA) has discussed concerns of a 2,150% increase in congenital syphilis in Oregon over the last 10 years, while other STIs are following national trends [15]. This finding highlights the importance of screening and treatment during pregnancy.

More patients received BPG for late latent syphilis in the post-intervention period than in the pre-intervention period, which was significantly different. When receiving treatment for late latent syphilis, the most common mistake was the use of only one injection. Appropriate treatment for late latent syphilis requires three BPG injections, each 1 week apart [14]. Factors such as patient loss to follow-up compounded the challenge of achieving proper late latent syphilis treatment. Since syphilis is a reportable condition, public health nurses would often communicate with patients or prescribe providers that additional doses of BPG are required for late latent syphilis. Other common reasons for inappropriate treatment include utilizing three BPG injections for recent exposures, primary infection, or early latent syphilis, instead of the one injection recommended [14]. Additionally, treatment may be based on syphilis serologies alone, without obtaining a sexual history, addressing syphilis treatment history, or recent exposure events. This illustrates the importance of proper sexual health history when deciding how to treat syphilis.

The majority of reactive RPR samples belonged to patients who serofast (pre-intervention, n = 146, 76.0%; and post-intervention, n = 157, 74.1%). Providers may be testing patients too early and not within the recommended intervals, leading to a high number of serofast samples. Most of the reactive RPRs also belonged to patients with a history of syphilis (pre-intervention n = 172, 89.6%, and post-intervention n = 184, 86.8%), emphasizing the importance of understanding the concept of serofast states and not overtreating.

A 58.1% increase in the use of patient education materials was another positive outcome. Notably, areas such as UC and the ED demonstrated the most consistent utilization of syphilis patient education. Patient education is a cornerstone of effective STI management as it enhances the understanding of disease transmission, treatment adherence, and the importance of partner notification. Providers can foster informed health behaviors and reduce reinfection rates by equipping patients with accessible and accurate information. Patient education equips individuals to manage the J-H reaction at home, forecast receiving a call from public health, and talk about re-exposure to syphilis when speaking to providers.

During the post-intervention period, views on the updated syphilis microlearning decreased. The availability of this tool needs to reach a broader audience to increase the use of practical resource microlearning. Some methods include presenting syphilis education tools during grand rounds of primary care to improve visibility. A discussion with a physician colleague who managed patient messages for the entire region revealed that the new syphilis resource was valuable and significantly improved, especially in understanding RPRs through infographics. The low usage of microlearning resources may be due to providers being comfortable treating syphilis, while others may use different resources, such as the University of Washington National STD Curriculum, the Centers for Disease Control and Prevention (CDC) STI Treatment Guide Mobile App, or clinical decision support tools, such as UpToDate. Emerging technologies, including artificial intelligence (AI)-driven tools that assist with RPR interpretation, also hold promise in supporting providers in complex diagnostic and treatment decisions, particularly in time-limited or high-volume care settings. Finally, since a third party provided the page view count, it is unclear whether the count reflects views per visit or per individual.

### Limitations

The implementation of education was uneventful, as there was buy-in from the stakeholders. However, engaging in a large, complex organization poses several challenges. Although regional notifications regarding syphilis patient education were announced via e-mail, the method of communication was new for the region, which may have led the nursing staff to overlook the notification or inadvertently delete it. Specific education provided to patients was missing from the documentation, and education was not attached to the patient’s AVS when the patient received BPG injections from the NTR.

Throughout the months following the intervention, outreach efforts were conducted with RN leads in three clinics, reminding the staff to incorporate syphilis education into their BPG administration. A future option is to attend meetings throughout the region to disseminate this information; however, at the time of implementation, this was not feasible. Finally, a limitation of this study was the lack of statistical significance in the dataset. Data were collected for only three months post-intervention, which affected statistical power. An extended observation period could result in statistically significant changes in educational and BPG administration habits.

## Conclusion

Utilizing patient education and clinician microlearning has revealed essential insights into how innovative educational strategies can improve syphilis treatment. Future steps include increasing the visibility of practice resource guidelines and automating patient education during visits for BPG injections. For example, automatically attaching education to a patient’s AVS and visiting note templates to include specific points for the RN to discuss with the patient. Adding short audiovisuals to syphilis microlearning can help clinicians develop diverse learning styles. Ultimately, with the increasing presence of AI in healthcare, there is an untapped opportunity to improve diagnostic accuracy, particularly in remote and underserved areas. Syphilis treatment can be complex; however, abbreviated sexual health history questions and understanding of the RPR titer can significantly improve treatment. This is especially important when it comes to people with prior syphilis infections who are in a serofast state.

## Supporting information

SQUIRE Checklist

## Data Availability

All data produced in the present work are contained in the manuscript

## Acknowledgements

Acknowledgement to individuals who made this project possible. Duke University: Nancy Crego, PhD RN, CHSE, FAAN; Mariam Kayle, PhD, RN, CCNS, FAAN; Leila Ledbetter, MLIS, AHIP; Elena Turner; Kaiser Permanente Northwest: Melisa Laurits-Stocek, MS, PA-C, AAHIVS; P. Alex Leahey, MD; Meagan Mangus, MSN, RN, NI-BC, CPHQ; Kelly Staten, MBA

## Ethical Compliance

The Kaiser Permanente Interregional IRB (KPiIRB) Office determined that this project did not meet the regulatory definition of research and did not involve human subjects

