## Supplemental Material for "Not So Fast, I’m Serofast: Using Innovative Education Techniques to Drive Management of People with Syphilis"

### Appendix #1: Key Stakeholder Interview

#### Patient affected by syphilis

- Got a call from the public health department
  - “I missed a lot of calls, thinking that it was spam calls.”
- Scheduling shots for syphilis was easy
  - “Walk-in” to get syphilis shot at Nurse Treatment Room
    - Knew that he had the option to schedule an appointment on his mobile app
- Did not know that he would have positive syphilis serologies for his lifetime
- Endorses that the shot was painful. However, he did not have any other side effects
- He did not receive any education on possible side effects and actions to be taken if this were to occur

*Registered Nurse*

- Did not realize there was a shortage of Benzathine Penicillin G (BPG)
- “I heard about patients remaining positive for syphilis for the rest of their lives”
- Often clinicians would order BPG with a diagnosis of “history of STI” but am unsure if BPG is indicated
- The nurse treatment room can see as many as 1-3 patients who need BPG injections a day and 1-3 BPG injections in a week.
- Nurses are good at providing education
  - Adding education to the “Wrap Up” activity in the EHR
  - Having 3 points to educate the patient on would be helpful
- It may be a good workflow to educate patients during the 15-minute monitoring post-BPG administering

Nurse Practitioner

- It is difficult to know which syphilis test to order
  - Syphilis reserve algorithm for people not previously infected with Syphilis versus just RPR titers for people who were previously infected
  - It would be “slick” to have the current EHR automatically order the correct test based on a patient’s history of syphilis infection
- Difficult and a lack of time to figure out if the patient had a history of syphilis and if treatment had been adequate
  - Often, patients don’t remember if they had syphilis or not
  - Did not realize that calling public health could be helpful
- It is difficult to understand and interpret RPR titers
- Speaking to clinicians in the urgent care setting may be helpful as they often treat and manage syphilis infections.

### Appendix #2: Search Table

| Database/Platform: Medline/PubMed  Date of Search: 2/12/2024 | | |
| --- | --- | --- |
| Search Set | Search Query | Results |
| #1: Syphilis | "treponema pallidum"[MeSH Terms] OR ("treponema"[All Fields] AND "pallidum"[All Fields]) OR "treponema pallidum"[All Fields] OR "syphilis"[All Fields] OR ("chancre"[MeSH Terms] OR "chancre"[All Fields] OR "chancres"[All Fields]) OR ("sexually transmitted diseases"[MeSH Terms] OR ("sexually"[All Fields] AND "transmitted"[All Fields] AND "diseases"[All Fields]) OR "sexually transmitted diseases"[All Fields]) OR "sexually transmitted infection"[All Fields] OR "sexually transmitted infections"[All Fields] | 438,878 |
| #2: Treatment | "penicillin g benzathine"[MeSH Terms] OR "penicillin g benzathine"[All Fields] OR ("penicillin g"[MeSH Terms] OR "penicillin g"[All Fields]) OR ("therapeutical"[All Fields] OR "therapeutically"[All Fields] OR "therapeuticals"[All Fields] OR "therapeutics"[MeSH Terms] OR "therapeutics"[All Fields] OR "therapeutic"[All Fields]) OR ("treatment outcome"[MeSH Terms] OR ("treatment"[All Fields] AND "outcome"[All Fields]) OR "treatment outcome"[All Fields]) OR ("therapeutics"[MeSH Terms] OR "therapeutics"[All Fields] OR "therapies"[All Fields] OR "therapy"[MeSH Subheading] OR "therapy"[All Fields] OR "therapy s"[All Fields] OR "therapys"[All Fields]) | 12,213,057 |
| #3: Patient Education | "patient education as topic"[MeSH Terms] OR ("patient"[All Fields] AND "education"[All Fields] AND "topic"[All Fields]) OR "patient education as topic"[All Fields] OR "patient education handout"[Publication Type] OR "patient education handout"[All Fields] | 113,481 |
|  | #1 AND #2 AND #3 | 2,927 |
|  | Filter: Clinical Trial, Meta-Analysis, Randomized Controlled Trial, Review, Systematic Review, Year 2014-2024 | 141 |

| *Database/Platform: CINAHL/EBSCOhost*  Date of Search: 2/12/2024 | | |
| --- | --- | --- |
| Search Set | Search Query | Results |
| #1: Syphilis | (MH "Syphilis, Congenital") OR (MH "Syphilis+") OR "syphilis" | 6,938 |
| #2: Treatment | (MH "Treatment Outcomes+") OR (MH "Treatment Errors+") OR (MH "Treatment Complications, Delayed") | 496,459 |
| #3: Patient Education | (MH "Patient Education+") OR "Patient Education" OR (MH "Patient Discharge Education") OR (MH "Electronic Health Records+") | 120,495 |
|  | #1 AND #2 AND #3 | 5 |

### Appendix #3: Clinician Microlearning

#### Pre-Intervention – Syphilis Screening Practice Resource

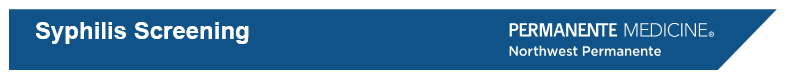

SUMMARY

Knowing whether the patient has a history of syphilis:

1. Determines which test you order
2. Determines how you interpret the results

To determine whether your patient has had syphilis in the past:

I. Check the medical record e.g. problem list and lab results (including Health Information Exchange)

2. Ask the patient

Syphilis testing should be part of comprehensive screening for other sexually transmitted infections, including HIV screening for other sexually transmitted infections, including HIV

ORDERING THE CORRECT SYPHILIS TEST

Type "syphilis" as an order.

Two choices appear:

- RPR NON-SCREENING, NO ALGORITHM
- SYPHILIS SCREENING ALGORITHM (lgG with reflex to RPR)

**Which one should you order?**

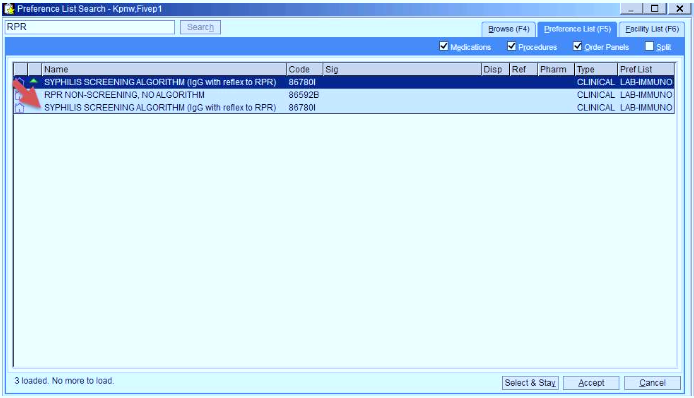

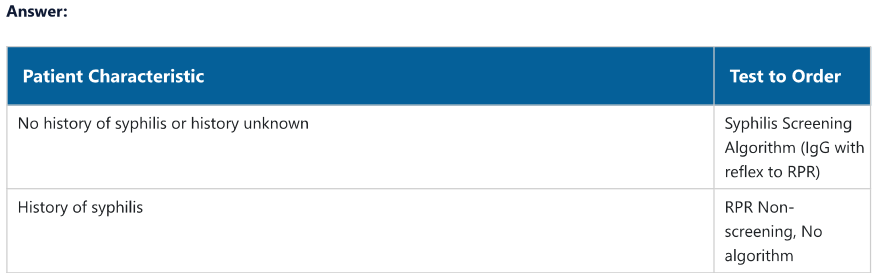

HOW DO I INTERPRET THE SYPHILIS SCREENING ALGORITHM?

If the Treponema pallidum lgG EIA screening test is negative — then the test for syphilis is negative.

If the Treponema pallidum lgG EIA screening test is positive — then the RPR will automatically be run.

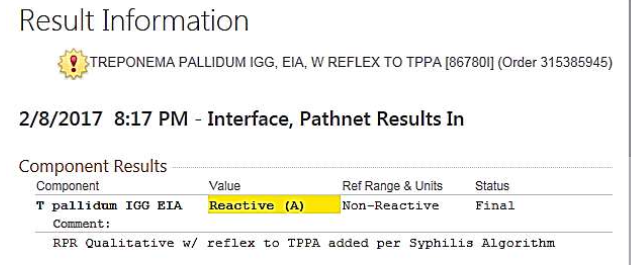

If both tests are positive, then either:

1. The patient has syphilis now, or
2. The patient had syphilis in the past but their RPR never became non-reactive
   - Also called "serofast"
   - Are usually low titers e.g. 1:1 to 1:4 range

If the T pallidum lgG is positive and the RPR is negative, then a 3rd "tie-breaker" test is run called the Treponema pallidum Antibody — Particle Agglutination (TPPA) test. That test result will be accompanied by the following narrative

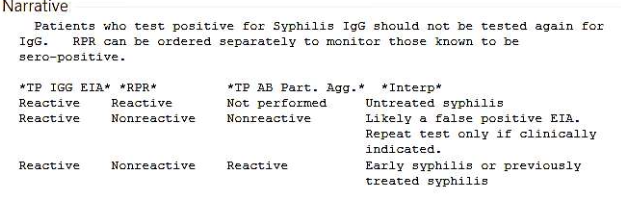

Interpretation of test results and action:

| **Treponema Pallidum IgG EIA (TP IGG EIA)** | **RPR** | **Treponema pallidum Antibody – Particle Agglutination (TP AB Part. Agg.)** | **Interpretation (Interp)** | **Action** |
| --- | --- | --- | --- | --- |
| Reactive | Reactive | Not performed | Untreated syphilis. | Treat.* |
| Reactive | Nonreactive | Nonreactive | Likely false positive EIA. Patients does not have syphilis. | Don't treat. |
|  |  |  | Uncommon - early syphilis, before RPR has had a chance to turn positive. If clinically indicated repeat the test after 2-4 weeks. | If the patient is high risk or has a painless chancre, then treat* and repeat the test. |
| Reactive | Nonreactive | Reactive | Previously treated syphilis. | If previously treated, don’t treat. If patient denies or doesn’t recall a history of treatment, then treat for late latent syphilis.* |
|  |  |  | Uncommonly- early syphilis, before RPR has had a chance to turn positive. If clinically indicated repeat the test after 2-4 weeks. | Don't treat. |
| * **Syphilis Treatment Practice Resource Page** | | | | |

RPR: INTERPRETING POSITIVE RESULTS IN PATIENTS WITH PRIOR HISTORY OF SYPHILIS

Basic principles:

1. A four-fold drop in titer within 12 months indicates successful treatment (e.g. 1:256 to 1:64 or less) for most patients

• Lower initial titers (1:8 or less), later syphilis stage, and older patients may be less likely to decline fourfold

2. Patients successfully treated, may have lingering low level positive results e.g. 1:1 or 1:2. This is called "serofast"

TREATMENT AND FOLLOW UP TESTING

**Link to Syphilis Treatment Practice Resource Page**

WHO SHOULD BE SCREENED?

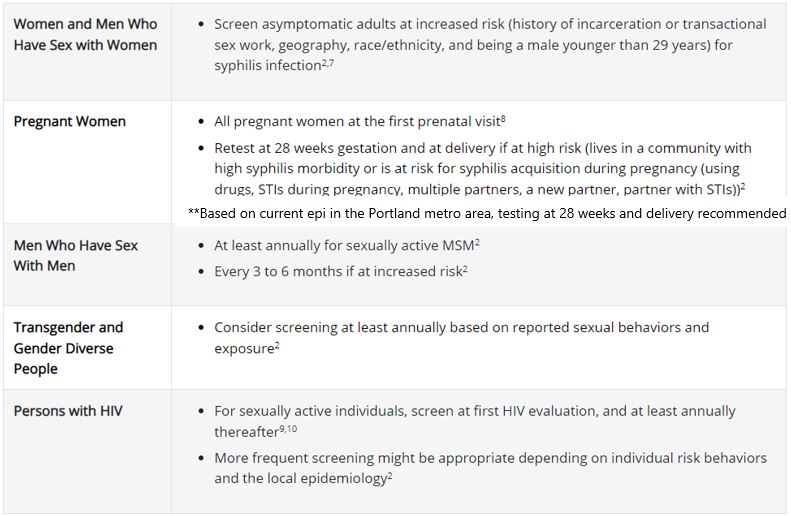

DEFINITIONS

Lab tests:

Non-specific (usually reactivity wanes over time)

- RPR (rapid plasma reagin)
- VDRL (Venereal Disease Research Lab)

Specific (usually remain reactive lifelong)

- T pallidum lgG EIA (enzyme immunoassay)
- TPPA (treponemal particle agglutination assay)
- FTA (fluorescent treponemal antibody)

Traditional screening algorithm

Screen with non-specific RPR, confirm with treponemal specific assay For example: RPR --> FTA

"Reverse" screening algorithm (our current testing method)

Screen with treponemal assay, then reflex to non-specific assay

At KPNW: T pallidum lgG EIA--> RPR --> T pallidum particle agglutination

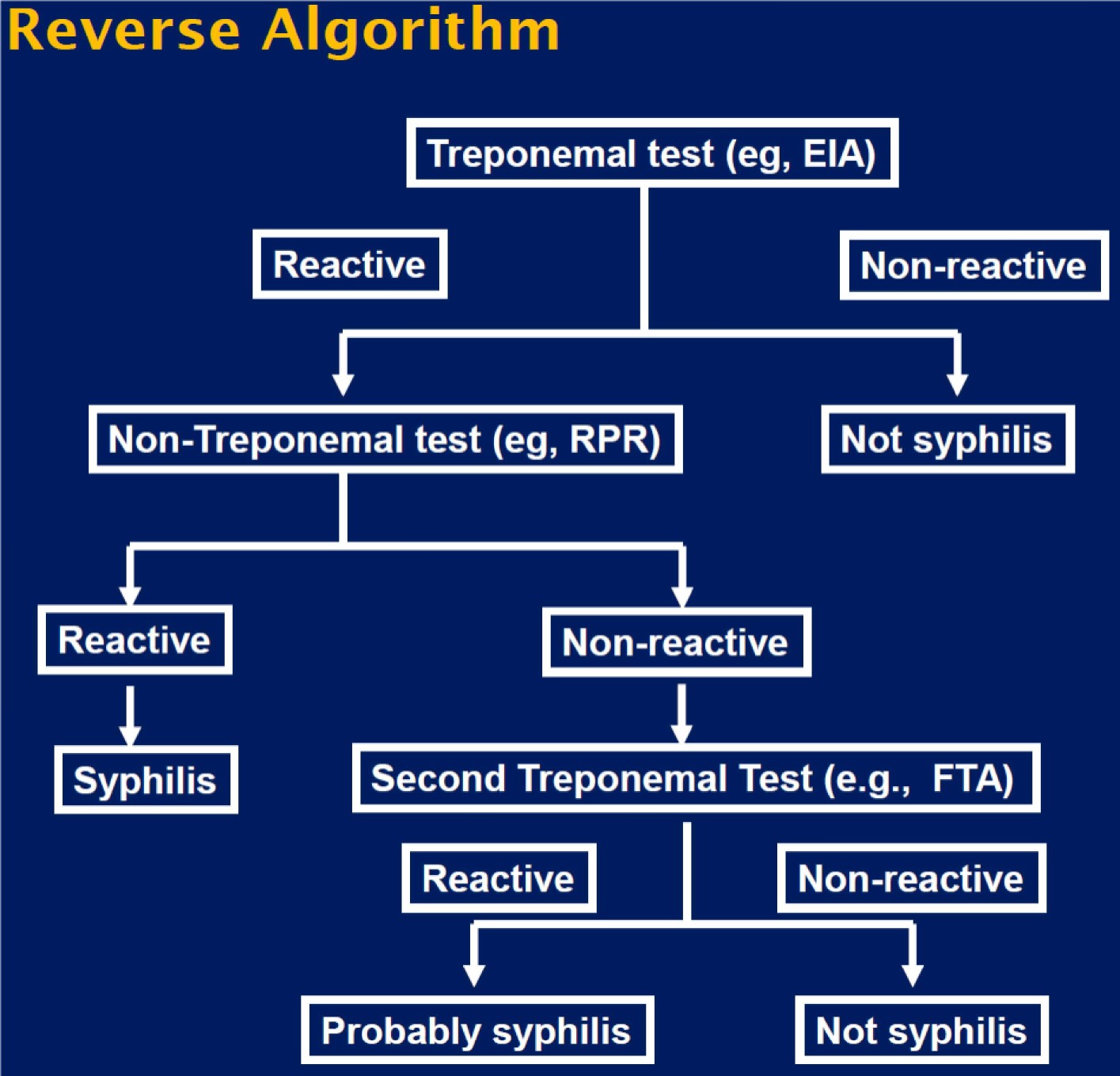

From Sean Schafer, MD, MPH. Oregon Public Health Division

DISCLAIMER

This guideline is informational. It is not a substitute for the reasonable exercise of independent clinical judgment by practitioners.  Each patient’s needs should be considered on an individual basis when rendering treatment plans. Recommendations are designed to apply to populations, not individuals.

© *2022 Northwest Permanente, P.C.; used with permission*

#### Post-Intervention – Syphilis Screening Practice Resource

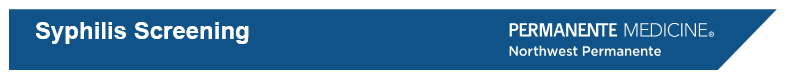

SUMMARY

Knowing whether the patient has a history of syphilis:

1. Determines which test you order
   - If there is a documented history of syphilis within the Electronic Health Record, RPR will be the default choice when ordering a syphilis test
2. Determines how you interpret the results

To determine whether your patient has had syphilis in the past:

1. Check the medical record e.g. problem list and lab results (including Health Information Exchange)
2. Ask the patient
3. Contact public health

Serofast refers to patients with persistent low titers (around 1:2-1:6) after having a ≥4-fold decrease, they do not need to be retreated.

Syphilis testing should be part of comprehensive screening for other sexually transmitted infections, including HIV.

ORDERING THE CORRECT SYPHILIS TEST

For people without a history of syphilis or whose history is unknown, type “syphilis” in the order panel and select “SYPHILIS SCREENING ALGORITHM” (#O388943)*

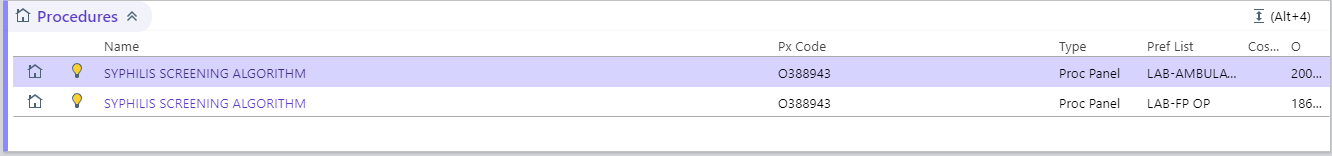

* If the patient has a documented history of syphilis within KP HealthConnect, “RPR W Titer” will be the default choice instead of “Syphilis Screening Algorithm”

For people with a history of syphilis outside of KP, type “RPR” in the order panel and select “RPR NON-SCREENING, NO ALGORITHM” (#253709)

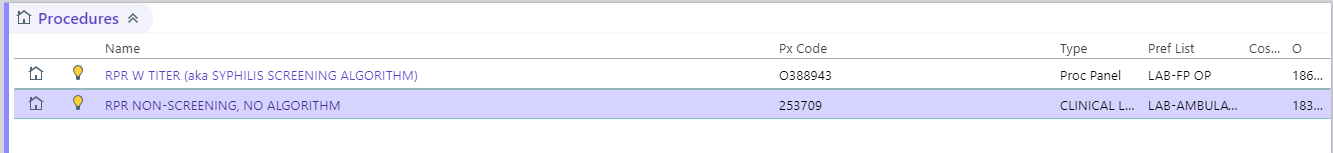

HOW DO I INTERPRET THE SYPHILIS SCREENING ALGORITHM?

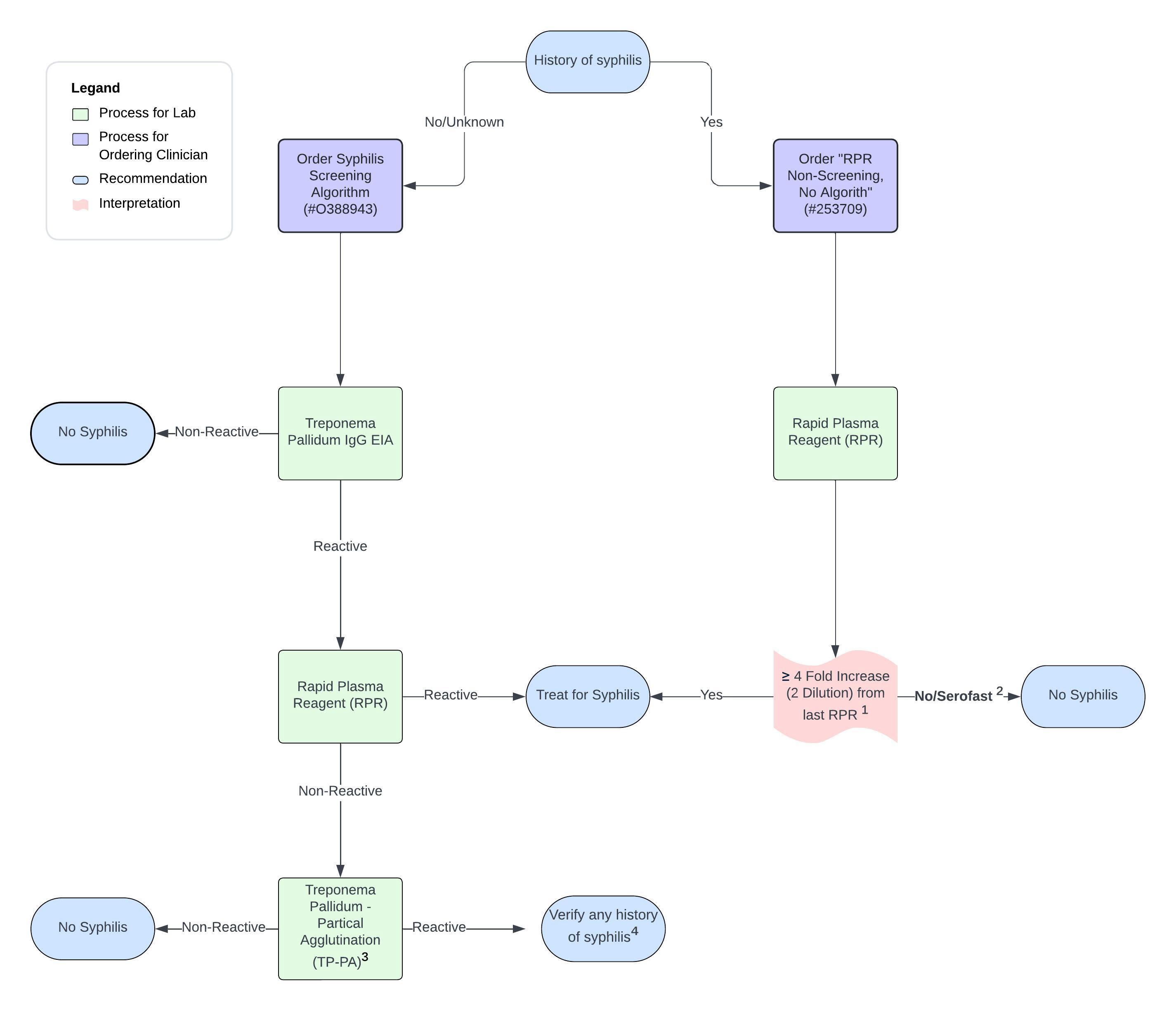

¹Example of ≥4-fold decrease is 1:128 to 1:32 of 1:256 to 1:64

²Serofast refers to patients who do not serorevert to and have persistent low titers (around 1:2-1:6) after having a ≥4-fold decrease

³TP-PA is used as a “tie breaker” when an EIA is reactive and RPR is non-reactive

Note: People with a history of syphilis will likely have a reactive treponema test for their lifetime

⁴Positive EIA and TP-PA with a negative RPR are likely from past syphilis infection. Verify/re-interview patients if they have a history of syphilis. Treatment using Bicillin within the indication can cause harm to the patient. If you have questions, please contact Public Health Communicable Disease Division or the HIV Prevention and Care (HPC) Clinician on Duty (COD)

Interpretation of test results and action:

| **Treponema Pallidum IgG EIA (TP IGG EIA)** | **RPR** | **Treponema pallidum Antibody – Particle Agglutination (TP AB Part. Agg.)** | **Interpretation (Interp)** | **Action** |
| --- | --- | --- | --- | --- |
| Reactive | Reactive | Not performed | Untreated syphilis. | Treat.* |
| Reactive | Nonreactive | Nonreactive | Likely false positive EIA. Patients does not have syphilis. | Don't treat. |
|  |  |  | Uncommon - early syphilis, before RPR has had a chance to turn positive. If clinically indicated repeat the test after 2-4 weeks. | If the patient is high risk or has a painless chancre, then treat* and repeat the test. |
| Reactive | Nonreactive | Reactive | Previously treated syphilis. | If previously treated, don’t treat. If patient denies or doesn’t recall a history of treatment, then treat for late latent syphilis.* |
|  |  |  | Uncommonly- early syphilis, before RPR has had a chance to turn positive. If clinically indicated repeat the test after 2-4 weeks. | Don't treat. |
| * **Syphilis Treatment Practice Resource Page** | | | | |

How do I interpret an RPR for a patient with a history of Syphilis

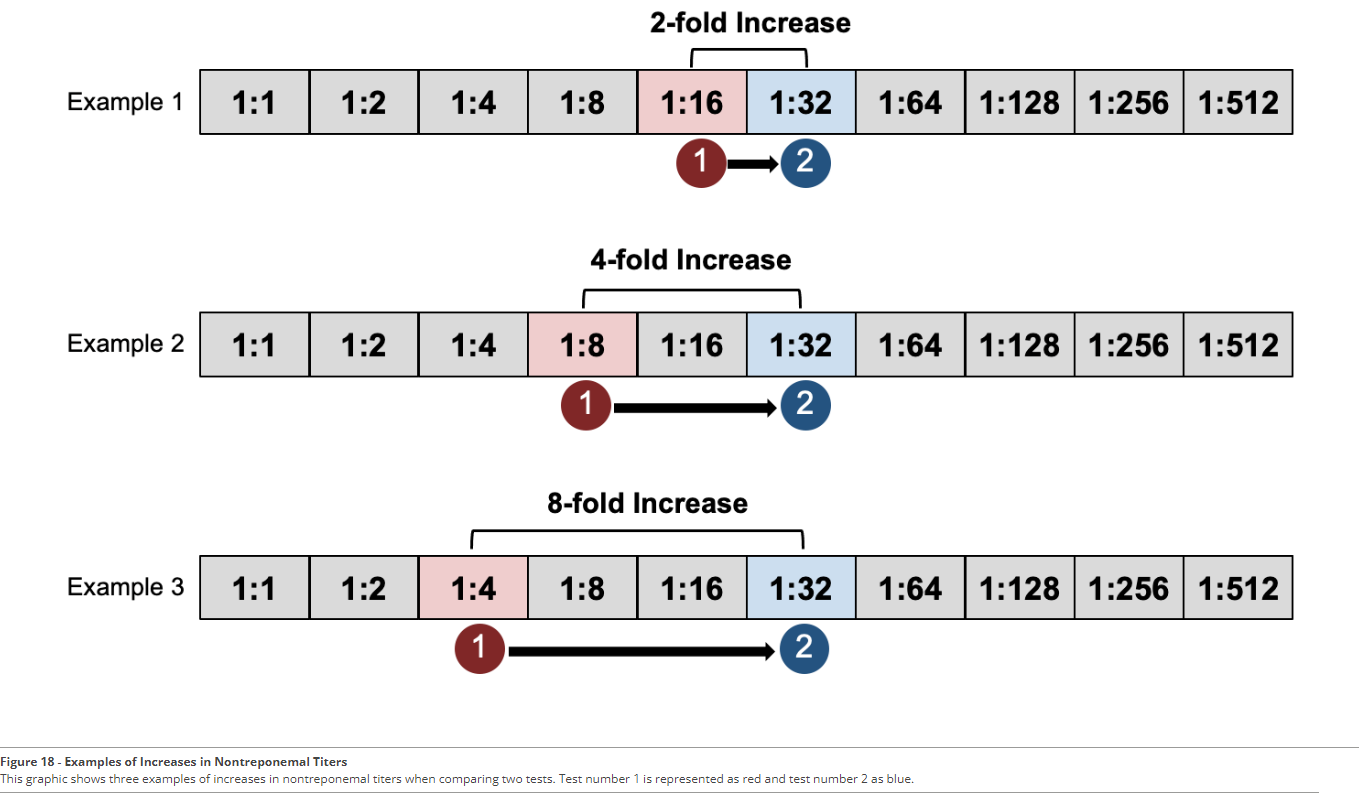

Criteria for successful treatment?

1. A four-fold drop in titer within 12 months indicates successful treatment (e.g. 1:256 to 1:64 or less) for most patients
   - Lower initial titers (1:8 or less), later syphilis stage, and older patients may be less likely to decline fourfold
   - Ensure that you are re-testing in the recommended interval, as retesting too early can lead to inaccurate results.
2. Patients successfully treated may have lingering low-level positive results e.g. 1:1 or 1:2. This is called “serofast”

My patient had a 4-fold (2 dilution) decrease in RPR, however that have lingering low titers such as 1:1 or 1:2 and not revert to non-reactive

- Some people will have adequate response who do not serorevert
  - This is referred to as “serofast”
- People who are serofast DO NOT require treatment, as long as there is not a 4-fold increase
- People who are serofast are likely to serorevert back to non-reactive RPR after two years [8, 18]

I do not know if my patient has had syphilis in the past.

- Interview patients and ask if they have been treated for syphilis in the past
- Public Health Communicable Diseases can have a good resource for assessing for prior syphilis tests and treatment
  - Patient resides in:
    - Multnomah: 503-988-3406
    - Clackamas: 503-655-8411
    - Clark: 564.397.8082
    - Washington: 503-846-3594
    - Marion: 503-588-8621
    - Yamhill: 503-434-7525

TREATMENT AND FOLLOW UP TESTING

**Link to Syphilis Treatment Practice Resource Page**

WHO SHOULD BE SCREENED?

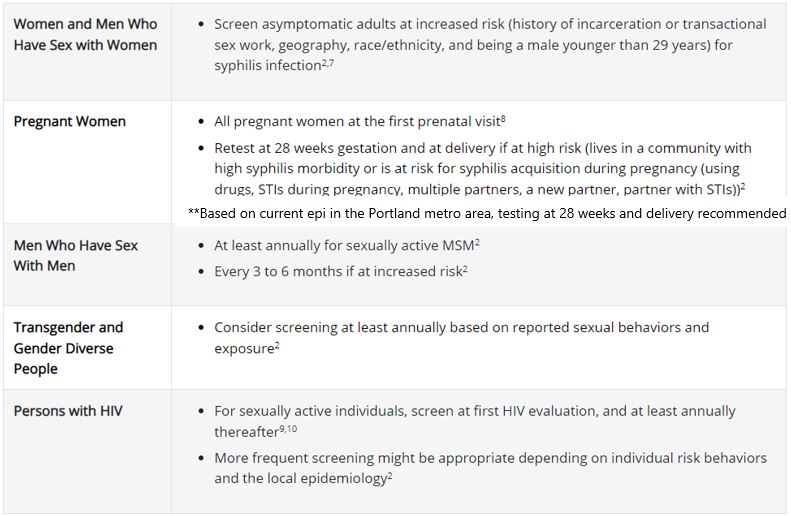

DEFINITIONS

Non-treponema test is an indirect method of measuring disease activity. Used for people with a history of syphilis

- Rapid Plasma Reagent (RPR)
- Venereal Disease Research Lab (VDRL)

Treponema test is a direct method of measuring antibodies. Likely remain reactive lifelong

- Treponema Pallidum IgG Enzyme immunoassay (EIA)
- Treponema Pallidum - Partial Agglutination (TP-PA)
- Fluorescent Treponemal Antibody (FTA)

**Contact HIV Prevention and Care (HPC) on-call for any additional questions or concerns about your patient’s syphilis result or treatment.**

DISCLAIMER

This guideline is informational. It is not a substitute for the reasonable exercise of independent clinical judgment by practitioners.  Each patient’s needs should be considered on an individual basis when rendering treatment plans. Recommendations are designed to apply to populations, not individuals.

© *2024 Northwest Permanente, P.C.; used with permission*

### Appendix #4: Patient Education

#### Pre-Intervention Patient Education

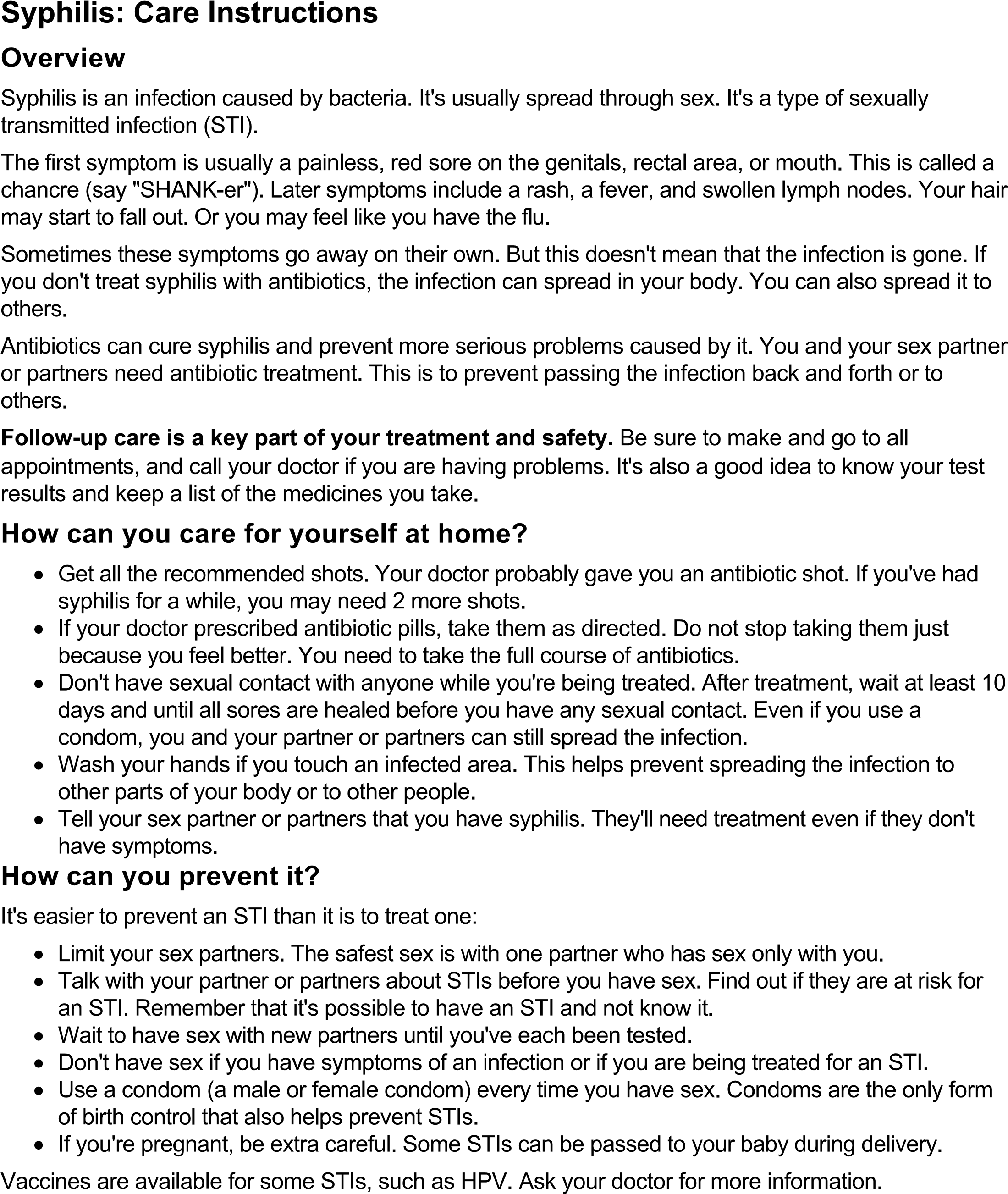

#### Post-Intervention Patient Education

**Syphilis**

**Your Care Instructions**

**Overview**

- Treatment for syphilis is with 1–3 weekly shots of an antibiotic called Bicillin, a penicillin.
- Common side effects include pain where the shot was given, rash, muscle pain, and fevers/chills within 24 hours after you receive treatment. This is not an allergic reaction.
- You will likely have a positive syphilis test for your lifetime. This doesn’t mean you still have syphilis.
- If you are re-exposed to syphilis, please contact your healthcare clinician to let them know you may have been re-exposed to syphilis and have had a positive test in the past. They will need to order a specific syphilis test called the rapid plasma reagin (RPR).

**What is syphilis?**

Syphilis is an infection you can get from having sex. Infections that are spread through sex are called "sexually transmitted infections" (STIs). It is also possible for a pregnant person to give syphilis to their baby.

The first symptom is usually a painless, red sore on the genitals, rectal area, or inside mouth. This is called a chancre ("SHANK-er"). If untreated, later symptoms may include a rash, a fever, swollen lymph nodes, and flu-like symptoms. Your hair may start to fall out.

Sometimes, these symptoms go away on their own. But this doesn't mean that the infection is gone. If you don't treat syphilis with antibiotics, the infection can spread in your body. You can also spread it to others.

**Treatment**

Antibiotics can cure syphilis and prevent more serious problems caused by untreated infection. You and your sex partners need treatment. This is to prevent passing the infection back and forth or to others.

The most common antibiotic treatment is an injectable type of penicillin called Bicillin. Your clinician will probably prescribe this for you.

You can make an appointment to get this shot at the Kaiser Permanente nurse treatment room that is most convenient for you.

Depending on how long you have had the syphilis infection, you may need 3 weekly doses of Bicillin to be sure of a cure. If your clinician prescribes multiple doses, it is important to complete all of them.

Often, people will have a fever and flu-like symptoms (headache, body aches) within 24 hours of antibiotic treatment for syphilis. This is called a Jarisch-Herxheimer (“yar-ish herks-hy-mer]) reaction. It is the body’s response to the syphilis bacteria dying. It is NOT an allergic reaction to penicillin. The reaction generally only lasts for a day. You can take ibuprofen, naproxen, or acetaminophen to help with the symptoms.

Do not be alarmed if you receive a phone call from your county's public health department. Their aim is to be sure you have been treated correctly, provide prevention education, and prevent further transmission.

**How can you care for yourself at home?**

- If your clinician prescribes antibiotic pills, take them as directed. Do not stop taking them just because you feel better. You need to take the full course of antibiotics.
- If you have the Jarisch-Herxheimer reaction, you can take ibuprofen, naproxen, or acetaminophen to help with the symptoms.
- During treatment, do not have sex. After completing treatment, wait for at least 7 days before having condomless sex.
- Tell your sex partners that you have been treated for syphilis. They need to be tested, even if they don't have symptoms.

**How can you prevent it?**

- Limit your sex partners. The safest sex is with one partner who has sex only with you.
- Talk with your partners about STIs before you have sex. Find out if they are at risk for any. Remember that it's possible to have an STI and not know it.
- Wait to have sex with new partners until you've each been tested.
- Don't have sex if you have symptoms of an infection or if you are being treated for an STI.
- Condoms (male and female condoms) are a very effective way to prevent STIs, particularly when used every time you have sex.
- If you're pregnant, be extra careful. Some STIs can be passed to your baby during delivery.
- If you are re-exposed to syphilis, please contact your health care clinician to let them know you may have been re-exposed to syphilis and have had a positive test in the past. They will need to order a specific syphilis test called the rapid plasma reagin (RPR).

Vaccines are available for some STIs, such as HPV. Ask your clinician for more information.

**When should you call for help?**

Watch closely for changes in your health. Be sure to contact your clinician if:

- You do not get better as expected.
- Your symptoms continue or come back after treatment.
- You develop new symptoms, such as a fever.

**Where can you learn more?**

Go to [About Syphilis | Syphilis | CDC](https://www.cdc.gov/syphilis/about/index.html)

### Appendix #5: Syphilis Staging

Syphilis Stage based on patient presentation and/or serology

| **Syphilis Staging** | **Definition** |
| --- | --- |
| Primary | Chancre or “primary lesion” noted [16] |
| Secondary | Diffused Maculopapular rash (particularly on palms of hands and soles of feet) noted.  Can also accompanied with fever, headache, malaise, anorexia, and/or diffuse lymphadenopathy |
| Early Latent | ≥ 4-fold increase RPR with the last known Syphilis Serology **≤ 12 months ago** without primary or secondary syphilis clinical manifestations |
| Late Latent | ≥ 4-fold increase RPR with the last known Syphilis Serology **> 12 months ago or Unknown** without primary or secondary syphilis clinical manifestations (Tuddenham & Ghanem, 2022) |
| Serofast | Persistent Low Syphilis Titers  **AND** ≥ 4-fold decrease in RPR titer post-treatment  **AND** absence of symptoms |
| Treatment <12 months ago for P&S or Early Latent | < 4-fold decrease in RPR titer in a **12-month**s period without a ≥4-fold increase in RPR  **AND** adequate post-treatment for primary, secondary, or early latent syphilis  **AND** absence of symptoms |
| Treatment <24 months ago for Late Latent | < 4-fold decrease in RPR titer in a **24-months** period without a ≥4-fold increase in RPR  **AND** adequate post-treatment for late latent syphilis  **AND** absence of symptoms |
| Exposure | Recent/known exposure event to syphilis |
| Congenital Syphilis | Vertical transmission of syphilis from a pregnant person to an infant |
| No Syphilis | No signs and symptoms of syphilis, recent/known exposure event and syphilis RPR and TP-PA serologies where non-reactive |

Centers for Disease Control and Prevention. Tertiary syphilis - STI treatment guidelines, https://www.cdc.gov/std/treatment-guidelines/tertiary-syphilis.htm (2021, accessed 10 March 2024).

Tuddenham S, Ghanem KG. Management of adult syphilis: Key questions to inform the 2021 Centers for Disease Control and Prevention sexually transmitted infections treatment guidelines. *Clinical Infectious Diseases* 2022; 74: S127–S133.

Fiumara N. Serologic responses to treatment of 128 patients with late latent syphilis. *Sex Transm Dis* 1979; 6: 243–246.
